# Climate change and environmental influence on visceral leishmaniasis in West Pokot county, Kenya

**DOI:** 10.1101/2022.05.08.22274805

**Authors:** Bulle Abdullahi, Joshua Mutiso, Fredrick Maloba, John Macharia, Mark Riongoita, Michael Gicheru

## Abstract

Kala**-**azar is a parasitic disease caused by *Leishmania species* transmitted by sand fly. In Kenya, kala-azar is endemic in thirty sub-counties spread over in eleven counties in the arid zones. Climate change-influenced seasonal weather variability and environmental alterations remain important determinants of many vector-borne diseases. The present study focused on climate change and environmental influence on kala-azar in West Pokot. Descriptive cross-sectional and retrospective research design was adapted. Study area was purposively selected. Locations and villages were randomly selected, while households were systematically selected. Three hundred sixty three household questionnaires, eleven key informant interviews, and five focus group discussions were undertaken. Secondary data was obtained from Kacheliba sub-county hospital records. Statistical package for social science version 24 was used to analyze quantitative data while NVivo software was used to analyse qualitative data. Kala**-**azar cases have been on the rise on aggregate from 2010 to 2020, 400 to 500 annual average cases, and incident rate tends to surge during dry season and just after the rains when humidity is high and temperature variations are wide. Significant environmental characteristics associated with risk of kala-azar infection included: presences of seasonal rain water pathways and rock piles around houses (X^2^= 30.706, df=1, p<0.001); presence of acacia trees in and around homesteads (X^2^=21.404, df=1, p<0.001); presence of dumping site (X^2^=9.641, df=1, p=0.002); presence of anthills around the homesteads (X^2^=16.538, df=1, p<0.001); presence of animal shed (X^2^=4.290, df=1, p=0.038); presences of chicken shelter (X^2^=36.862, df=1, p<0.001); and practices of frequently moving houses to new temporary compound (X^2^=20.919, df=1, p<0.001). Climate change-induced seasonal weather variability; increased temperature and reduced precipitation and environmental alterations remain significant predictors of kala-azar in West Pokot. Community sensitization on disease prevalence, clearing of vector predilection sites and improving community environmental risk perception are imperative to promote prevention and reduce disease burden.

**Brief summary:** Climate change and environmental alteration influence on vector-borne diseases is getting stronger as ever increasing global temperatures and human activity-induced environmental changes remain key drivers of vector expansion and disease re-emergence. In Kenya, kala-azar cases are on an increasing trend as annual incidents surpassed 1,500 with a growing list of over eleven endemic counties. The present study used household questionnaire, interrogated surrounding environment using observation checklist and delved into Kacheliba hospital records. Risk of kala**-**azar was significant in association with presence of seasonal water pathways, presence of acacia trees, anthills, cattle & goat and chicken shelters around houses. On aggregate, kala-azar incident rate was increasing from 2010 to 2020, and cases tend to surge towards end of first and start of last quarter annually. These surge periods coincide with dry season and just after rains in the area when the humidity is high. Apparently during the last 10 years there seems reduced precipitation and increase temperature. Interestingly, increased in temperature and reduced precipitation was associated with increased reported Kala-zar cases. Policy makers and concerned agencies should consider promoting preventive behaviors, increasing community risk perception and eliminating vector-harboring structures around houses, while observing environmental conservation as a disease mitigation strategy.

## 1. Introduction

Leishmaniasis is a vector borne protozoan re-emerging disease caused by *Leishmania species* and transmitted by sand fly vectors [1,2]. Leishmaniasis is a neglected disease receiving little development and research investments, thus, considered as a disease of poverty as majority of its victim’s are people with humble economical background who reside at the periphery of global villages where basic resources like health are limited [3,4]. Leishmaniasis has four major disease forms which range from simple skin lesions to fatal systemic infection and include: cutaneous form, visceral form, muco**-**cutaneous form, and post kala-azar dermal leishmaniasis form [5,6]. Visceral leishmaniasis is considered most lethal among all the forms with over 95% fatality rate if untreated and second most killer parasitic disease after malaria [4,7,8]. Although only 25**-**45% of the incidents are reported, visceral leishmaniasis is endemic in over hundred countries worldwide and more than ninety percent of the cases occur in ten countries mainly in Eastern Africa, South-East Asia region, and Brazil [4]. Globally, visceral leishmaniasis is causing more than 50,000 fatalities, over one million cases, and more than 350 million people are at risk [9,10].

After the Indian sub-continent, Eastern Africa is considered to be contributing second most highest number of visceral leishmaniasis cases and they are mainly caused by *Leishmania donovani* [11,12]. In Kenya, visceral leishmaniasis was first documented in early 19^th^ century when an outbreak occurred around Lake Turkana area [13]. Currently in Kenya, visceral leishmaniasis is endemic in over eleven counties which are reporting approximately 1,500 cases annually, and more than five million people are at risk of this disease mostly in the arid and semi-arid climatic zone [5,14,15]. Considering the fast setting-in of climate change effect which triggers frequent alterations on seasonal weather pattern, both the global prevalence and emergence of vector borne diseases in uncommon areas is expected to rise [16]. Regardless of *phlebotomine species* involved and reservoir behaviors, weather fluctuations and environmental modifications are considered strong determinants of leishmaniasis [17,18]. Similarly, global warming and climate change-induced alteration in temperature and precipitations variability remains to be significant predictors in vector epidemiology in terms of abundances and activeness [19]. In African where more than seventy percent of the population lives in rural areas, it is expected that the continent will experience more of the effect of climate change and global warming as a result of the emerging new settlements and widespread alteration of environment through deforestation which may promote creation of more vector breeding sites [20]. Increase in temperature is believed to favor sand fly multiplication while also increasing its period of activity [21]. Environmental factors surrounding homesteads including presences of certain plant species, seasonal rain water pathways-created land fissure and crevices, presences of animal sheds, and chicken houses are thought to influence the presence as well as activity of the vector [2,18].

The devastating effect of global warming and climate change coupled with prevalent low individual immune system and social weakness in the visceral leishmaniasis endemic zones, cases of leishmaniasis are expected to get aggravated, thus, the need to undertake epidemiological studies particularly in endemic areas [22–24]. The study aimed to contribute to the sustainable development goal number 3.3, World Health Organization’s neglected tropical diseases road map target 2021 to 2030, as well as Kenya’s ministry of health 2021**-**2025 strategic plan for control of visceral leishmaniasis and vision 2030. The present study endeavors to ascertain climate change and environmental influence on visceral leishmaniasis in West Pokot county in order to provide essential information to help in up-scaling strategies to reduce visceral leishmaniasis burden.

## 2. Materials and methods

### Study area

The current study was carried out from September 2020 to February 2021 in West Pokot county and particularly in Pokot north sub-county. The sub**-**county has 3,910 km^2^ land mass and is home to 134,485 persons [25]. The area experiences temperature of 20 ^0^c to 39^0^c and mostly seasonal rainfall of approximately 500 mm. Pokot north borders with Turkana county to the north and east and international border with Uganda to the west.

### Study design

The study combined both descriptive cross**-**sectional and retrospective study design aimed at establishing climate change and environmental alterations**-**induced seasonal weather variability influence on visceral leishmaniasis burden from 2010 to 2020 in West Pokot county.

### Sampling

The study area was selected purposively considering the high number of kala**-**azar cases reported and presence of visceral leishmaniasis treatment and diagnostic centre in the study area. Random sampling of locations and villages, and systematic selection of household that lived in the area for more than six months was undertaken. Three hundred sixty three household questionnaires and observation checklist, eleven key informant interviews, and five focus group discussions were carried out as well as secondary data obtained from Kacheliba hospital records. Historical Kacheliba temperature and rainfall amount obtained from world weather online was used as weather parameter. Sample size was determined by using Cochran’s n=Z^2^Pq/e^2^.

### Data analysis

Statistical Package for the Social Sciences (SPSS) version 24 and Microsoft excel software were used to analyse quantitative data, and chi**-**squire was used to test for the level of association between variables. Qualitative data was analysed using NVivo software to establish connection and interpretation in identification of patterns. Correlation analysis between the disease burden and temperature or rainfall amount was done using Pearson correlation test. Only variables with P-value of less than 0.05 were considered significant.

## 3. Results

### 3.1 Environmental characteristics of household respondents

Table 1 describes distribution of environmental characteristics that were considered significant in influencing the risk of visceral leishmaniasis in the study area. More than half (53.1%) of the respondents had seasonal rain water pathways or rock piles existing around their homesteads while 46.9% had none of these in close proximity. Almost three quarters of the participants (74.7%) had acacia trees present in and around their compounds while about a quarter (25.7%) had either uprooted or lived in areas where this tree species was not common. A considerable percentage (37.5%) of respondents had garbage dumping sites present within fifty meters proximity to their houses, while the majority (62.5%) reported to dispose waste haphazardly in their farm fields and surrounding community rangeland fields. Majority (81.7%) of the study participants had anthills present in and around their compounds, while few (18.3) had either removed them or naturally the anthills were not common in their area of residence. More than (80%) of the respondents had constructed animal sheds and chicken shelters within their compounds. Considering traditional practice of moving houses to temporary new compounds, small percentage (29.2%) of respondents reported to be practicing traditional nomadic habit of frequently moving houses from time to time and to different locations.

**Table 1:** Household respondence to environmental characteristics in Pokot north sub-county of West Pokot County, Kenya (N=360)

| Variable | Category | n | % |
| --- | --- | --- | --- |
| Seasonal water pathways/rock pile presence | Yes | 191 | 53.1 |
|  | No | 169 | 46.9 |
| Acacia tree presence | Yes | 269 | 74.7 |
|  | No | 91 | 25.3 |
| Dumping site presence | Yes | 135 | 37.5 |
|  | No | 225 | 62.5 |
| Anthill presence | Yes | 294 | 81.7 |
|  | No | 66 | 18.3 |
| Animal shed presence | Yes | 291 | 80.8 |
|  | No | 69 | 19.2 |
| Chicken shelter presence | Yes | 318 | 87.8 |
|  | No | 40 | 12.2 |
| Recently moved house | Yes | 105 | 29.2 |
|  | No | 255 | 70.8 |

### 3.2 Household respondent environmental characteristics and association with visceral leishmaniasis

Cross-tabulation result in Table 2 on environmental characteristics and visceral leishmaniasis prevalence indicate that risk was high 33% (n=63) among those homesteads near seasonal rain water pathways and rock piles, and among those having acacia trees 27.5% (n=74) within their compounds. Similarly, higher occurrence of visceral leishmaniasis was observed among those households with dumping sites present 30.4% (n=41) approximately within fifty meters to the houses, and among those with presence of anthills 25.9% (n=76) in and around their compounds. The disease was also more prevalent among those households with constructed animal sheds 24.1% (n=70), and chicken shelters 17% (n=53) in and around their compound. Significant percentage 37.1% (n=39) among those who practiced moving houses from time to time to new temporary compounds also appeared to be at higher risk of visceral leishmaniasis compared to those who lived in permanent compounds 15.3% (n=39).

**Table 2:**
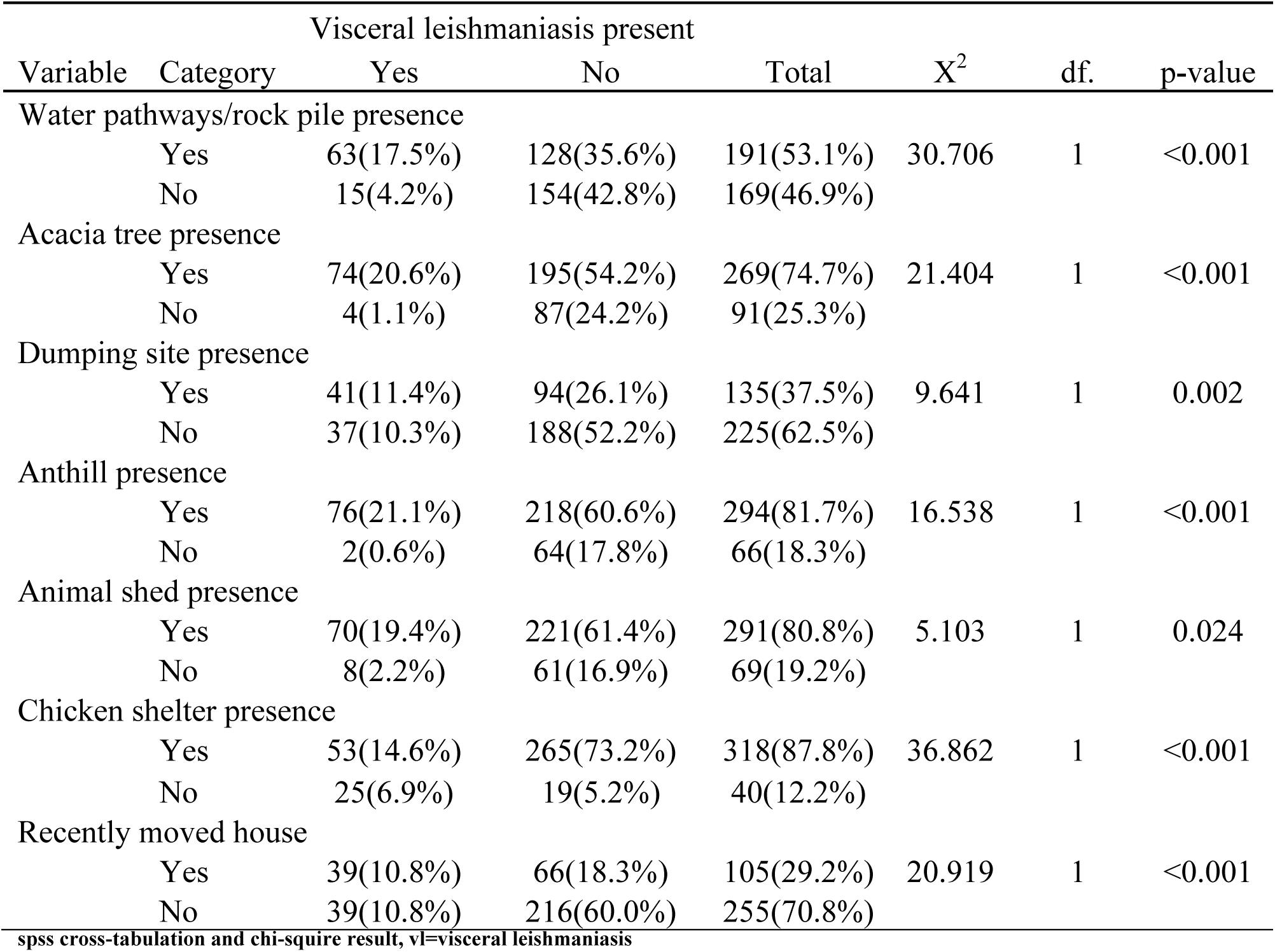
Household respondent environmental characteristics and association with visceral leishmaniasis in Pokot north sub-county of West Pokot county, Kenya (n=360)

The result in Table 2 further indicates that there is statistically significant association between risk of visceral leishmaniasis among respondents and presence of seasonal rain water pathway and rock piles around the compound (X^2^= 30.706, df=1, p<0.001). Presence of acacia trees in the compound and surroundings was also significantly associated with higher risk of infection (X^2^= 21.40, df=1, p<0.001). Presence of garbage dumping site approximately fifty meters to the house was statistically associated with increased risk of exposure to disease (X^2^= 9.641, df=1, p**<**0.002). Existence of anthills in and around the compound was also significant in relation to risk of visceral leishmaniasis infection (X^2^=16.538, df=1, p<0.001). Presences of constructed animal sheds and chicken shelters around the houses was significantly associated with risk of infection (X^2^=5.103, df=1, p=0.024) and (X^2^=36.862, df=1, p<0.001) respectively. Risk of visceral leishmaniasis was also significant among those who practice nomadic habit of moving houses to new temporary compounds and locations from time to time (X^2^=20.919, df=1, p<0.001; Table 2).

### 3.3 Trends of kala-azar incident rate and temperature from the year 2010 to 2020 in Kacheliba sub-county

Figure 1 shows annual visceral leishmaniasis incident rates as obtained from Kacheliba sub-county hospital records and Kacheliba sub-county annual temperature trends as obtained from World weather online (https://www.worldweatheronline.com/kacheliba-weather/rift-valley/ke.aspx) for the years 2010 to 2020. The result shows overall increase in average daily temperature from 23 ^0^c in 2010 to 26 ^0^c in 2020 (annually aggregate increase from 8169 ^0^c in 2010 to 9100 ^0^c in 2020). Similarly, the annual kala-azar incident rates in Kacheliba sub**-**county hospital tends to be increasing on aggregate from approximately 400 cases to almost 500 cases. Regarding the relationship, the results shows that annual kala**-**azar incident rate and annual temperature trends tend to have weak positive correlation (r=0.609; P=0.47).

**Figure 1:**
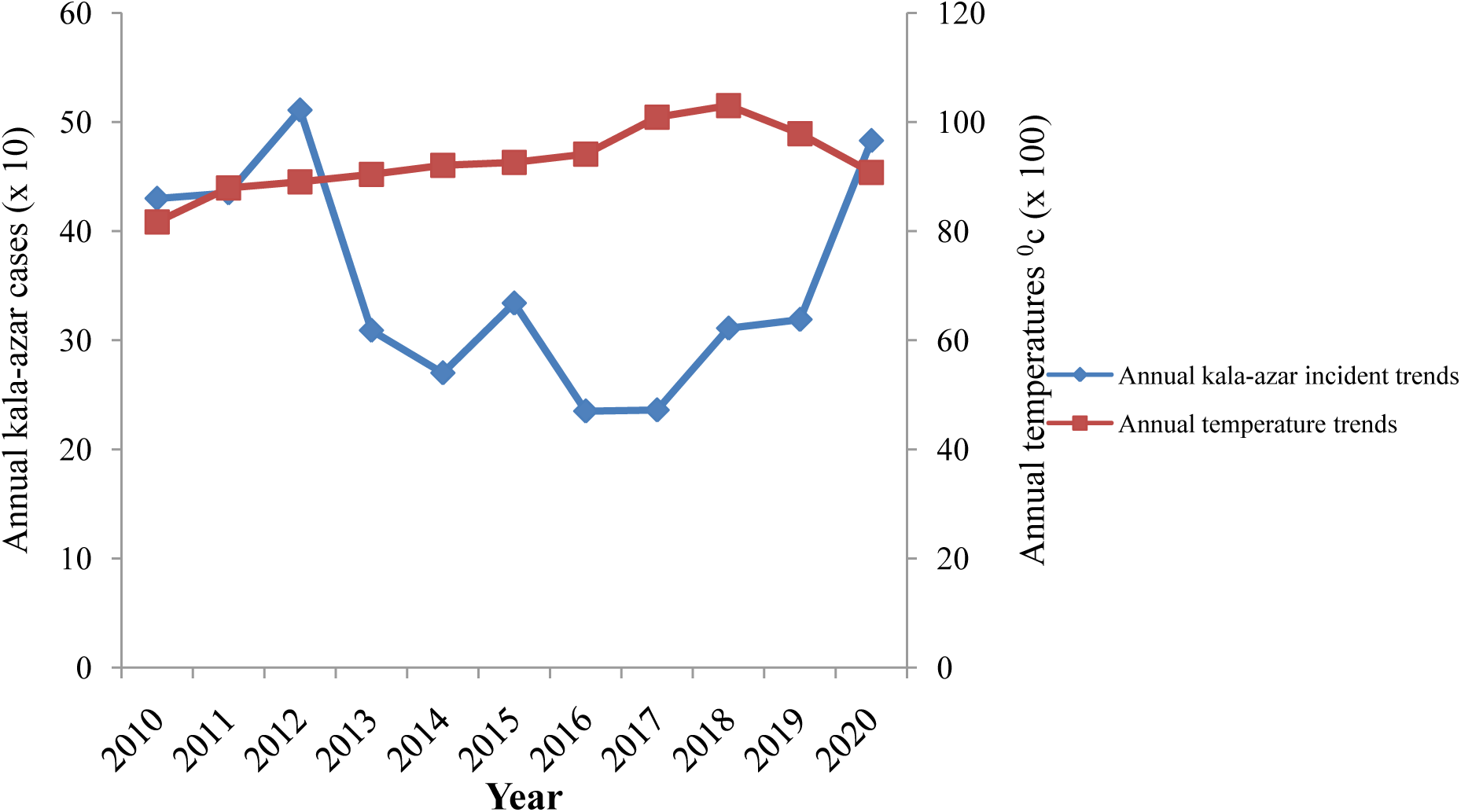
Trends of kala-azar incident rates and temperature from 2010 to 2020 in Kacheliba sub-county.

### 3.4 Trends of kala-azar incident rate and rainfall amount experienced in Kacheliba sub-county from 2010 to 2020

The result in figure 2 depicts the level at which the amount of rainfall received in Kacheliba sub**-**county had been diminishing over the last ten years from 2010 to 2020 as obtained from world weather online (https://www.worldweatheronline.com/kacheliba-weather/rift-valley/ke.aspx). The result indicates the overall annual rainfall amount in Kacheliba sub-county had reduced from 3643 mm in 2010 to 2976 mm in 2020, while the aggregate annual kala**-**azar incident rates over this decade increased from about 400 to approximately 500 cases annually. Precipitation in Kacheliba sub-county tends to have significant negative correlation with annual kala**-**azar incident rates (r= - 0.7; P=0.016).

**Figure 2:**
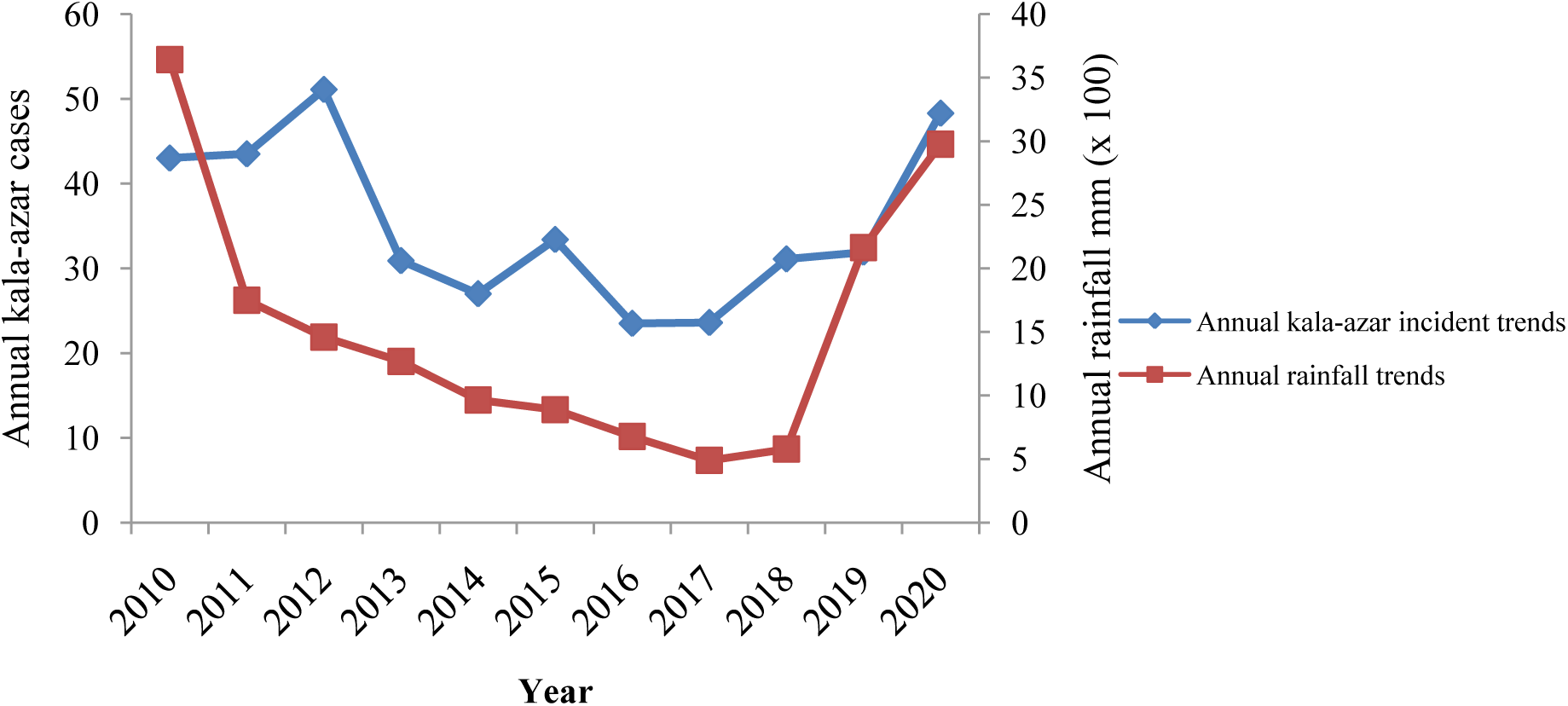
Trends of kala-azar incident rates and rainfall in Kacheliba sub-county from 2010 to 2020.

### 3.4 Seasonal variation of visceral leishmaniasis incidents in Kacheliba sub-county

Figure 3 shows the number of visceral leishmaniasis cases treated at Kacheliba hospital per month during the last half a decade from 2016 to 2020 years as obtained from Kacheliba sub-county hospital records. According to the hospital records, cases have been increasing from approximately three hundred in 2016 to slightly under five hundred in 2020. Visceral leishmaniasis cases per month ranged between twenty and approximately thirty during this half a decade. Concerning seasonal influence, visceral leishmaniasis incidence tend to be high during end of first quarter around March and April as well as start of last quarter around October and November every year, while cases remain relatively low during middle quarter of the years.

**Figure 3:**
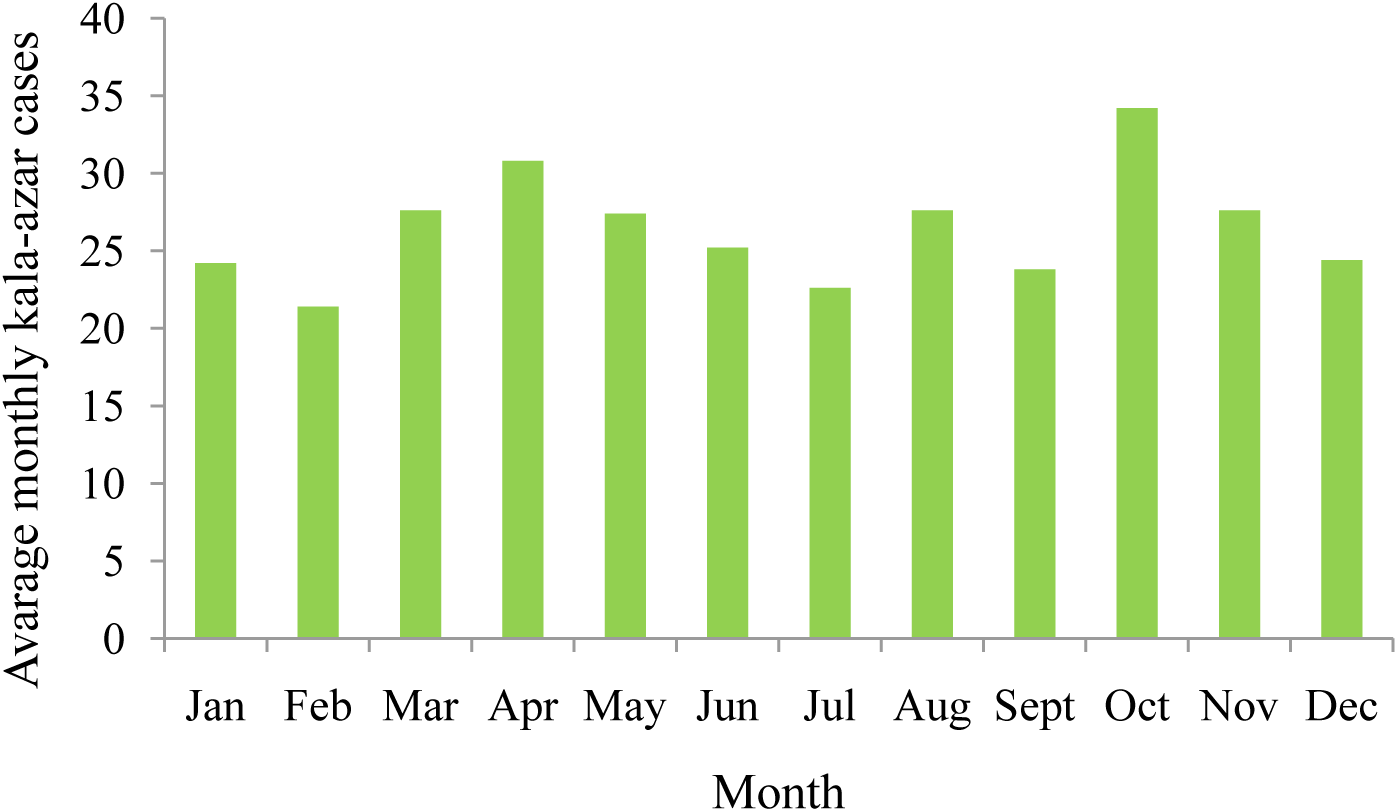
Seasonal variations of kala-azar incident rates from 2016 to 2020as obtained from Kacheliba sub-county hospital records.

## 4. Discussion

Visceral leishmaniasis is endemic in Kacheliba sub**-**county and remains to be the second most prevalent parasitic disease after malaria, and these findings agree with previous reports [3,7]. The present findings point out a multiplex relationship between kala**-**azar incidents and climate change, as an increase in temperatures and decrease in rainfall amount remain significant predicators of kala**-**azar incident rate in Kacheliba from 2010 to 2020. Moreover, the observed surges in kala-azar incident rates during dry season as well as just after the rains while remaining relatively low during cold weather winter season depicts the level of interaction between climate change**-**induced seasonal weather variability and their impact on kala-azar. The increase in kala**-** azar cases during the dry period is further attributed to elevated vector population as well as vector expansion to uncommon areas towards the end of dry seasons which is usually characterized by high humidity and wider temperature variations, and these findings support previous reports [21,27]. Similarly, climate change induced**-**prolonged customary drought conditions result in depletion of pasture coverage and triggers mass movement of humans and animals into deeper virgin high vector density rangeland sections in search of pasture and browsing content for their livestock which in the process exposes people to vector bite, and these findings support previous reports [28–31].

The significantly associated environmental factors; existence of seasonal rain water channels, presence of acacia tree, garbage dumping sites and anthills in and around human houses with high kala**-**azar infection rates may indicate the complexity of factors predisposing the communities to disease. Ever green big trees on the seasonal riverbanks provide shades from scotching sun to both herders and their livestock during dry period, likewise these trees as well as riverbank fissure and crevices form humid vector preferred hiding and breeding zones, hence providing risky areas of interaction [32,2,18]. Moreover, during flooding period these seasonal rain water pathways tend to move the sand fly larvae from the endemic zones to a new different places, hence increasing risk of transmission as well as vector expansion to new areas [33]. Due to the presence of disposed organic waste content among other characteristics, garbage dumping sites are frequented by reservoirs like rodents and disease transmitting vectors including sand fly, thus, their presence around homesteads attract and harbour the vector in the endemic areas [35,34]. The moderately humid atmosphere and plenty of hiding areas inside the anthill holes is believed to form perfect resting and breeding environment for the sand fly, thus, its presences around the human houses increases risk of infections [36,2]. The flowers and secretions of the acacia tree provide both feed and water to the sand fly, while it’s bark provides perfect resting and breeding place, hence presence of acacia in and around the compound poses risk of attracting the vector and exposing those living nearby [37,36,2].

Presence of animal sheds and chicken shelter as well as movement of houses from time to time to new temporary compounds and locations was associated with risk of visceral leishmaniasis infection, and this pose complication in disease control as these practices are cultural in nature for these communities. Many times animal and chicken shelters tend to be wet long after rains, thus, creating moisture and humidity that are believed to attract the vector while the structure provides resting spaces [38,39]. Traditional nomadic style of frequently moving houses and people to new compounds which is commonly practiced by the pastoral community living in the vector endemic arid and semi-arid areas is associated with high risk of infection. The risk is believed to be arising from the high vegetation density in the newly settled temporary compound which may be already invested with the vector or attract it as a result of human activities such as newly cultivated agricultural fields [40,41,30]. Our data is inconsistent with our earlier report by Macharia (2018) in Baringo, Kenya in which the community could not associate risk of kala-azar infection and environmental factors [42]. Interestingly, this may possibly exemplify the poor risk perception of the pastoral community on environmental associated risk factors of kala-azar infection. The limitations of present study remain to be those of cross**-**sectional and retrospective studies as data on household kala-azar incident rate and environmental risk factors were simultaneously assessed, as well as gaps of facility records data. In conclusion, the impact of environmental factors and climate change**-**induced seasonal weather pattern variability emerges as important determinant of vector distribution and habitats as well as increasing level of kala**-** azar burden in study area. The study recommends improvement in community risk perception, introduction of culturally acceptable behavior and habitat change, and reducing vector harboring structures around houses while observing environmental conservation to mitigate impact of climate change on kala-azar.

## Data Availability

All data produced in the present work are contained in the manuscript

## Ethical consideration

This study obtained approval from Kenyatta university ethical review committee [PKU/2067/I1214]. Clearance was obtained from West Pokot County department of health and informed consent sought from research participants.

## Data availability

All data collected and analyzed for this study are included in the article.

## Acknowledgments

The study acknowledges the residents of West Pokot county for their study participation, and community health volunteer Isaac Nyeris for his assistance in data collection. Kacheliba hospital medical superintendent Mr. Solomon Tukey and hospital administrator Ms. Melap Nakuya are credited for their support, as well as West Pokot county department of health for the research approval.

